# The impact of PEEP on hemodynamics, respiratory mechanics, and oxygenation of children with PARDS

**DOI:** 10.1101/2024.01.18.24301487

**Authors:** Fernanda Monteiro Diniz Junqueira, Isabel de Siqueira Ferraz, Fábio Joly Campos, Toshio Matsumoto, Marcelo Barciela Brandão, Roberto José Negrão Nogueira, Tiago Henrique de Souza

## Abstract

**Objective:** To assess the impact of increasing positive end-expiratory pressure (PEEP) on hemodynamics, respiratory system mechanics, and oxygenation in children with pediatric acute respiratory distress syndrome (PARDS).

**Design:** Prospective single-center study.

**Setting:** Tertiary care, university-affiliated PICU.

**Patients:** Mechanically ventilated children with PARDS.

**Interventions:** PEEP was sequentially changed to 5, 12, 10, 8, and again to 5 cmH_2_O. After 10 minutes at each PEEP level, hemodynamic and respiratory variables were registered. Aortic and pulmonary blood flows were assessed through transthoracic echocardiography, while respiratory system mechanics were measured using the least squares fitting method.

**Measurements and Main Results:** A total of 31 patients were included, with median age and weight of 6 months and 6.3 kg, respectively. The main reasons for PICU admission were respiratory failure caused by acute viral bronchiolitis (45%) and community-acquired pneumonia (32%). At enrollment, most patients had mild or moderate PARDS (45% and 42%, respectively), with a median oxygenation index of 8.4 (IQR 5.8–12.7). Oxygen saturation improved significantly when PEEP was increased. However, although no significant changes in blood pressure were observed, the median cardiac index at PEEP of 12 cmH_2_O was significantly lower than that observed at any other PEEP level (*p*=0.001). Fourteen participants (45%) experienced a reduction in cardiac index of more than 10% when PEEP was increased from 5 cmH_2_O to 12 cmH_2_O. Also, the estimated oxygen delivery was significantly lower at 12 cmH_2_O PEEP. Finally, respiratory system compliance significantly reduced when PEEP was increased. At a PEEP level of 12 cmH_2_O, static compliance suffered a median reduction of 25% (IQR 39.7–15.2) in relation to the initial assessment (PEEP of 5 cmH_2_O).

**Conclusions:** Despite the improvement in oxygen saturation, increasing PEEP in hemodynamically stable children with PARDS can cause a significant reduction in cardiac output, oxygen delivery, and respiratory system compliance.

**Key Points:**

- **Question:** What is the impact of positive end-expiratory pressure on hemodynamics, respiratory mechanics and oxygenation in children with acute respiratory distress syndrome?
- **Findings:** In this prospective single-center study, we found a significant reduction in stroke volume index and cardiac index when PEEP was increased to 12 cmH_2_O. Furthermore, despite the improvement in oxygenation, the increase in PEEP was associated with a significant reduction in the estimated oxygen delivery and respiratory system compliance.
- **Meaning:** In addition to oxygenation, PEEP titration in children should include close monitoring of hemodynamics and respiratory mechanics.

**RESEARCH IN CONTEXT:**

- Lung-protective ventilation using positive end-expiratory pressure (PEEP) remains the mainstay of respiratory management in ARDS.
- High PEEP levels have the potential to impact cardiac function and lung mechanics.
- Due to concerns about the adverse effects of high PEEP levels, hypoxemia is often managed by increasing the fraction of inspired oxygen rather than escalating PEEP.

**AT THE BEDSIDE:**

- Although it can improve peripheral oxygen saturation, high levels of PEEP have the potential to decrease cardiac output and thereby decrease oxygen delivery.
- As no changes in blood pressure were observed during PEEP titration, it cannot be used as a surrogate for cardiac output monitoring.
- Lung recruitability should be carefully evaluated in children with PARDS, as increasing PEEP may lead to reduced compliance of the respiratory system.

## INTRODUCTION

Pediatric acute respiratory distress syndrome (PARDS) is a life-threatening condition characterized by non-cardiogenic pulmonary edema and hypoxemia. Its incidence is around 3.2% of patients in PICU with mortality of up to 33% for severe cases (1, 2). While there is no specific therapy for PARDS, supportive therapies using lung-protective mechanical ventilation strategies have been associated with reduced mortality in adults and children (3, 4). Such strategies aim to reduce ventilator-induced lung injury by minimizing the strain and stress applied to the lung by mechanical ventilation. Thus, current PARDS guidelines recommend limiting plateau pressures (or driving pressures), low tidal volumes, high positive end-expiratory pressure (PEEP), and acceptance of permissive hypercapnia and some degree of hypoxemia.

Determining the optimal PEEP is one of the most challenging tasks in the management of PARDS. Optimal PEEP promotes adequate gas exchange with minimal end-expiratory atelectasis, alveolar overdistension, and hemodynamic impairment. Unfortunately, PEEP titration is often performed by only assessing oxygen saturation, which can lead to setting inappropriate PEEP levels. While insufficient PEEP can induce lung derecruitment and atelectrauma, excessive PEEP can cause alveolar overdistension, reduced venous return, and increased right ventricular afterload (5). Thus, despite the improvement in oxygenation, high PEEP levels can compromise respiratory mechanics and worsen oxygen delivery due to reduced cardiac output (6–9). Hence, the process of determining optimal PEEP comprises a comprehensive and simultaneous assessment of respiratory system compliance, hemodynamics and oxygen delivery.

Due to concerns about the adverse effects of high PEEP levels, pediatric intensivists often prefer to manage hypoxemia by increasing the fraction of inspired oxygen (FiO_2_) rather than escalating PEEP (10–12). Certainly, the lack of available evidence describing the effects of PEEP in the pediatric population contributes to the use of inappropriately low PEEP levels in children. Therefore, the main objective of the present study was to assess the impact of increasing PEEP on hemodynamics, respiratory system mechanics, and oxygenation in children with PARDS.

## METHODS

This prospective study was conducted at the PICU of the Clinical Hospital of the State University of Campinas (UNICAMP), Sao Paulo, Brazil. The study protocol was approved by the local institutional review board (UNICAMP’s Research and Ethics Committee; approval number: 28780820.4.0000.5404; approval date: October 13, 2021; Study title: Evaluation of the influence of positive end-expiratory pressure on hemodynamic variables and respiratory mechanics in children with acute respiratory distress syndrome) and written informed consent was obtained from the legal guardians of all participants. The procedures followed in this study were in accordance with the Helsinki Declaration of 1975. All procedures were followed according to the ethical standards of the local institutional review board and the Helsinki Declaration of 1975.

Mechanically ventilated children meeting PALICC PARDS criteria were assessed for eligibility (13). Although the study was conducted prior to publication of the PALICC-2, all participants also met the current PARDS definition (14). Inclusion criteria were as follows: 1) mechanical ventilation on pressure-controlled - volume guaranteed (PCV-VG) mode; 2) tidal volume of 6-8 ml/kg of predicted body weight; 3) leak around the endotracheal tube less than 20%; and 4) need for PEEP titration at the discretion of the attending physician. Participants were excluded if they met any of the following criteria:

1) clinical signs of respiratory efforts; 2) hemodynamic instability, defined as abnormal values of blood pressure or heart rate for age (15); 3) previously diagnosed congenital or acquired heart diseases; 4) suspected or confirmed pulmonary hypertension; 5) heart arrythmia; 6) chest wall deformities; and 7) need for titration of vasoactive drugs during the study period.

### Study Protocol

Right after enrollment, participants had their demographic and respiratory data collected. Then, PEEP was sequentially changed to 5, 12, 10, 8 and again to 5 cmH_2_O. Other ventilator settings remained unchanged throughout the whole study period, except for FiO_2_, which was adjusted to maintain oxygen saturation at 92-97%. After 10 minutes at each PEEP level, the following variables were evaluated:

- Hemodynamics: arterial blood pressure, heart rate, cardiac index (Ci), stroke volume index (SVi), aortic velocity-time integral (VTI), aortic blood flow peak velocity variation (ΔVpeak), pulmonary VTI, and pulmonary ΔVpeak;
- Respiratory mechanics: peak inspiratory pressure (PIP), mean airway pressure (MAP), inspiratory flow resistance, plateau pressure, auto-PEEP, static compliance (C_stat_), and dynamic compliance (C_dyn_);
- Oxygenation: FiO_2_, peripheral oxygen saturation (SpO_2_), oxygen saturation index (OSi), and estimated oxygen delivery index (eDO_2_);

For the analysis of the eDO_2_, the partial pressure of oxygen in the plasma was considered negligible and the hemoglobin concentration unchanged between the data collection periods. Thus, eDO_2_ was determined by the following formula: eDO (ml. min^-l^. m^-2^) = Ci x (1,34 x Hb x SpO2).

The body surface area of the participants was calculated using the Mosteller formula (16). All patients had their weight and height measured after admission to the PICU.

Aortic and pulmonary blood flows were assessed through transthoracic echocardiography (TTE) using an ultrasound machine (Vivid Q; GE Healthcare, Tirat Carmel, Israel) equipped with a phased array transducer (3.5–8 MHz). All echocardiographic examinations were performed by an experienced pediatric ultrasound instructor from the Brazilian Society of Intensive Care Medicine.

All patients were ventilated using a Hamilton-C1 ventilator (Hamilton Medical AG, Bonaduz, Switzerland). Tidal volume and respiratory rate remained unchanged throughout the study protocol. The Hamilton-C1 ventilator measures flows and tidal volumes proximal to the inspiratory limb. Respiratory mechanics is measured dynamically and continuously using the least squares fitting method (17). This method attempts to fit the equation of motion to the measured pressure, volume, and flow data to calculate respiratory mechanics (C_stat_, resistance, and auto-PEEP). Non-invasive blood pressure, heart rate and SpO2 were measured using Life Scope G5 monitors (CSM-1500; Nihon Kohden, Tokyo, Japan). Data obtained with the mechanical ventilator and the bedside monitor were recorded precisely after 10 minutes. No averages were taken for these variables during the evaluation period.

Details regarding the precision of every device utilized in this study can be found in

## Supplemental Digital Content

### Statistical analysis

Statistical analysis and sample size calculation were performed using MedCalc Statistical Software version 19.8 (MedCalc Software bvba, Ostend, Belgium) and G*Power software version 3.1.9.6, respectively (18). The normality of the data distribution was assessed using the Kolmogorov– Smirnov and Shapiro–Wilk tests. Continuous variables are expressed as medians and interquartile ranges (IQR), as data were not normally distributed. Categorical variables are expressed as absolute numbers and frequency (%). A *p*-value ≤ 0.05 was considered statistically significant. Since the data did not follow a normal distribution, differences between multiple observations of variables were analyzed using Friedman’s test. If a difference was found with Friedman’s test, then a post-hoc analysis was performed for pairwise comparison of variables according to Conover (19). Significance was defined as *p* < 0.05.

Due to the non-parametric distribution of the data, precise calculation of the sample size was not feasible. As an alternative, we computed the sample size for the parametric equivalent test and factored in an additional 15% as compensation (20). Conducting a power analysis for a repeated measures ANOVA involving five measures, with a power of 0.90, an alpha level of 0.05, and assuming a medium effect size (f = 0.25), revealed a required sample size of 26. After accounting for the nonparametric test by adding 15%, the necessary participant count became 30.

## RESULTS

Between October 2021 and December 2022, 35 patients were evaluated for eligibility. Three patients were excluded due to congenital heart disease and one due to pulmonary hypertension. Thus, 31 children were enrolled in the study, of which 17 (55%) were males. Demographic characteristics of the study population are presented in **Table 1**.

**Table 1.** Distribution of demographic and clinical characteristics of the participants (n = 31). Data are presented as median (interquartile range) unless otherwise specified.

| Variables | Value |
| --- | --- |
| Age (months) | 6.0 (3.0 – 12.7) |
| Weight (kg) | 6.3 (5.0 – 11.0) |
| Male sex, n (%) | 17 (55%) |
| Diagnosis, n (%) |  |
| - Acute viral bronchiolitis | 14 (45%) |
| - Community-acquired pneumonia | 10 (32%) |
| - COVID-19 | 4 (13%) |
| - Aspiration pneumonia | 3 (10%) |
| PIM-2 | 1.6 (0.32 – 5.4) |
| Oxygenation index | 8.4 (5.8 – 12.7) |
| pH | 7.38 (7.32 – 7.41) |
| PaO<sub>2</sub> (mmHg) | 84 (71 – 98) |
| PaCO<sub>2</sub> (mmHg) | 50 (45 – 61) |
| PARDS severity, n (%) |  |
| - Mild | 14 (45%) |
| - Moderate | 13 (42%) |
| - Severe | 4 (13%) |
| Mean airway pressure (cmH<sub>2</sub>O) | 13 (12 – 15) |
| Peak inspiratory pressure (cmH<sub>2</sub>O) | 29 (25-32) |
| Positive end-expiratory pressure (cmH<sub>2</sub>O) | 7 (5 – 8) |
| Auto positive end-expiratory pressure (cmH<sub>2</sub>O) | 0 (0 – 0.4) |
| FiO<sub>2</sub> (%) | 60 (40 – 70) |
| Delivered tidal volume (mL.kg<sup>-1</sup>) | 6 (5.5 – 6.5) |
| Respiratory rate (breath.min<sup>-1</sup>) | 28 (26 – 32) |
| Static compliance (mL.cmH<sub>2</sub>O<sup>-1</sup>.kg<sup>-1</sup>) | 0.38 (0.30 – 0.44) |
| Dynamic compliance (mL.cmH<sub>2</sub>O<sup>-1</sup>.kg<sup>-1</sup>) | 0.30 (0.21 – 0.38) |
| Inspiratory flow resistance (cmH<sub>2</sub>O.L<sup>-1</sup>.s<sup>-1</sup>) | 88 (58 – 140) |
| Plateau pressure (cmH<sub>2</sub>O) | 23 (19 – 27) |
| Driving pressure (cmH<sub>2</sub>O) | 16 (13 – 21) |

Abbreviations: PaO<sub>2</sub>, arterial oxygen partial pressure; PaCO<sub>2</sub>, partial pressure of arterial carbon dioxide; PIM, pediatric index of mortality; PARDS, pediatric acute respiratory distress syndrome; FiO<sub>2</sub>, inspired fraction of oxygen.

Some hemodynamic variables varied significantly with changes in PEEP levels (**Table 2**). Although no significant changes in blood pressure were observed, the median Ci at PEEP of 12 cmH_2_O was significantly lower than that observed at any other PEEP level (*p*=0.001). Initial values of SVi and aortic VTI were similar to those observed with PEEP of 8 and 5 cmH_2_O, but were significantly higher than those observed when PEEP was 10 or 12 cmH_2_O (*p*<0.001). Fourteen participants (45%) experienced a reduction in Ci and SVi of more than 10% when PEEP was increased from 5 cmH_2_O to 12 cmH_2_O. Regarding pulmonary blood flow, there was a significant reduction in the pulmonary VTI at PEEP levels of 12 and 10 cmH_2_O when compared to those measured at the beginning and end of the study protocol (PEEP of 5 cmH_2_O). A significant increase in aortic ΔVpeak was observed when applying PEEP of 12 cmH_2_O, but the same did not occur with pulmonary ΔVpeak. A complete pairwise analysis of all studied variables is presented in ***Supplemental Digital Content***.

**Table 2.**
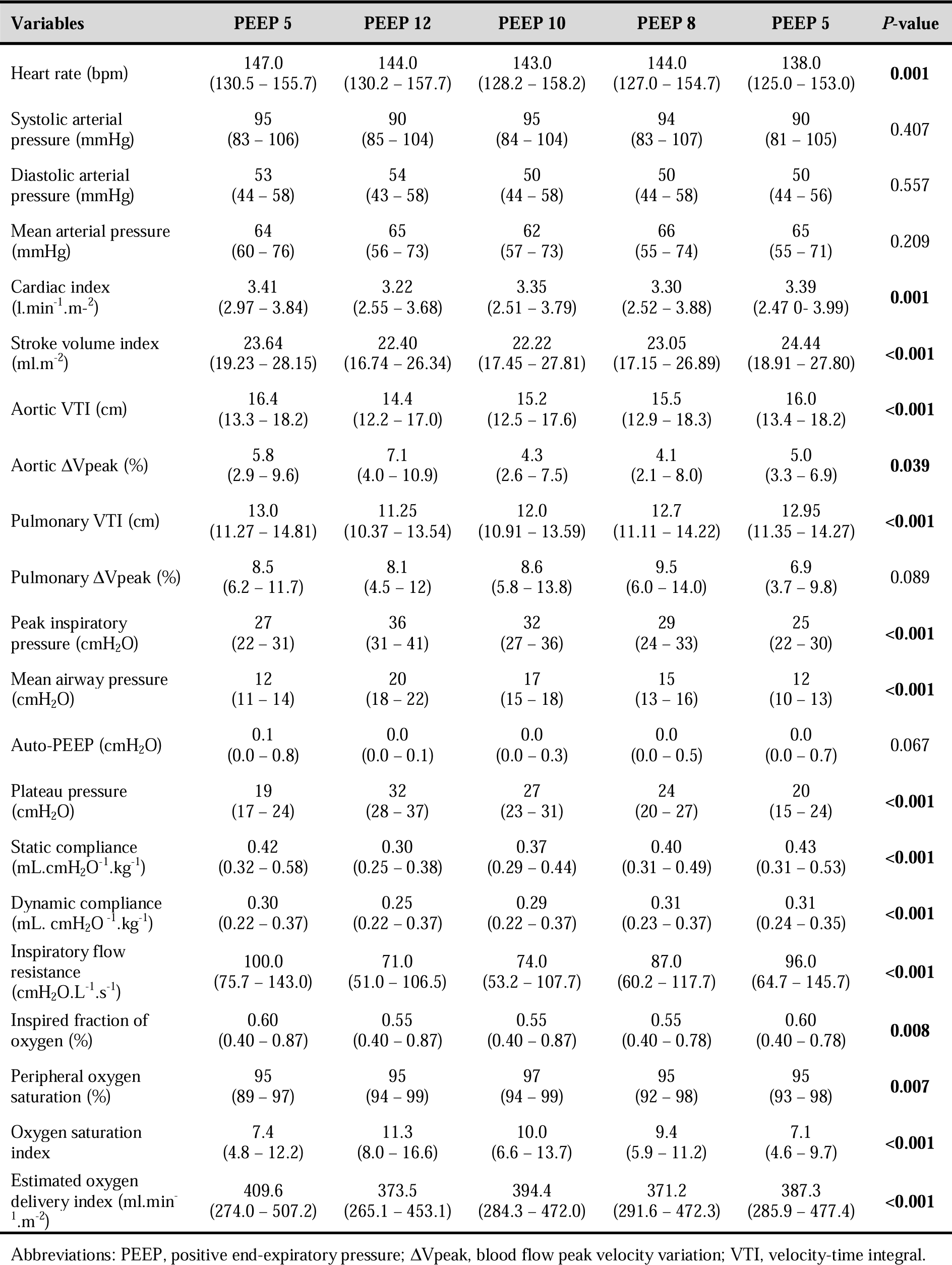
Hemodynamic, ventilatory and oxygenation variables at different levels of positive end-expiratory pressure. Differences between multiple observations of variables were analyzed using Friedman’s test. Post-hoc analysis for pairwise comparison of variables are presented in the Appendix. Values are expressed as median (25^th^–75 ^th^ percentiles).

With the exception of auto-PEEP, all other respiratory mechanics variables assessed showed significant changes during the study period (**Table 2**). Respiratory system compliance reduced significantly when PEEP was increased. Median values of C_stat_ were significantly lower at PEEP levels of 12 and 10 cmH_2_O than those observed with PEEP of 5 and 8 cmH_2_O (*p*<0.001). At a PEEP level of 12 cmH_2_O, C_stat_ suffered a median reduction of 25% (IQR 39.7 – 15.2) in relation to the initial assessment (PEEP of 5 cmH_2_O). Only two participants showed a 5% and 14% improvement in C_stat_ at this PEEP level. They were admitted due to COVID-19 and had mild and severe PARDS at the time of enrollment. At the end of the study protocol, the C_stat_ of 11 participants improved by at least 10%, while in 4 participants a reduction greater than 10% was observed in relation to the values collected in the initial phase (PEEP of 5 cmH_2_O). However, there was no significant change in the median C_stat_ value between these two evaluation moments (0.42 mL.cmH_2_O ^-1^.kg^-1^, IQR 0.32 – 0.58 vs 0.43 mL.cmH_2_O ^-1^.kg^-1^, IQR 0.31 – 0.53; *p*=0.162).

Median SpO_2_ values significantly increased when PEEP was changed from 5 to 12 cmH_2_O. This increase remained significant at PEEP of 10 cmH_2_O. However, the increase in SpO_2_ was greater than 10% of baseline values in only two participants, both with severe PARDS (OI of 23 and 24). Despite the improvement in SpO_2_, these two participants experienced a 16% and 39% reduction in C_stat_ with increasing PEEP, but there were no significant changes in their SVi. Although there was an increase in SpO_2_ when PEEP was scaled to 12 cmH_2_O, this did not lead to an improvement in the eDO_2_ (***Figure 1***). There was a significant reduction in eDO_2_ with increasing PEEP to 12 cmH_2_O. A reduction in eDO_2_ greater than 10% was observed in 14 patients (45%), while 10 patients (32%) experienced a reduction exceeding 15%.

**Fig. 1.**
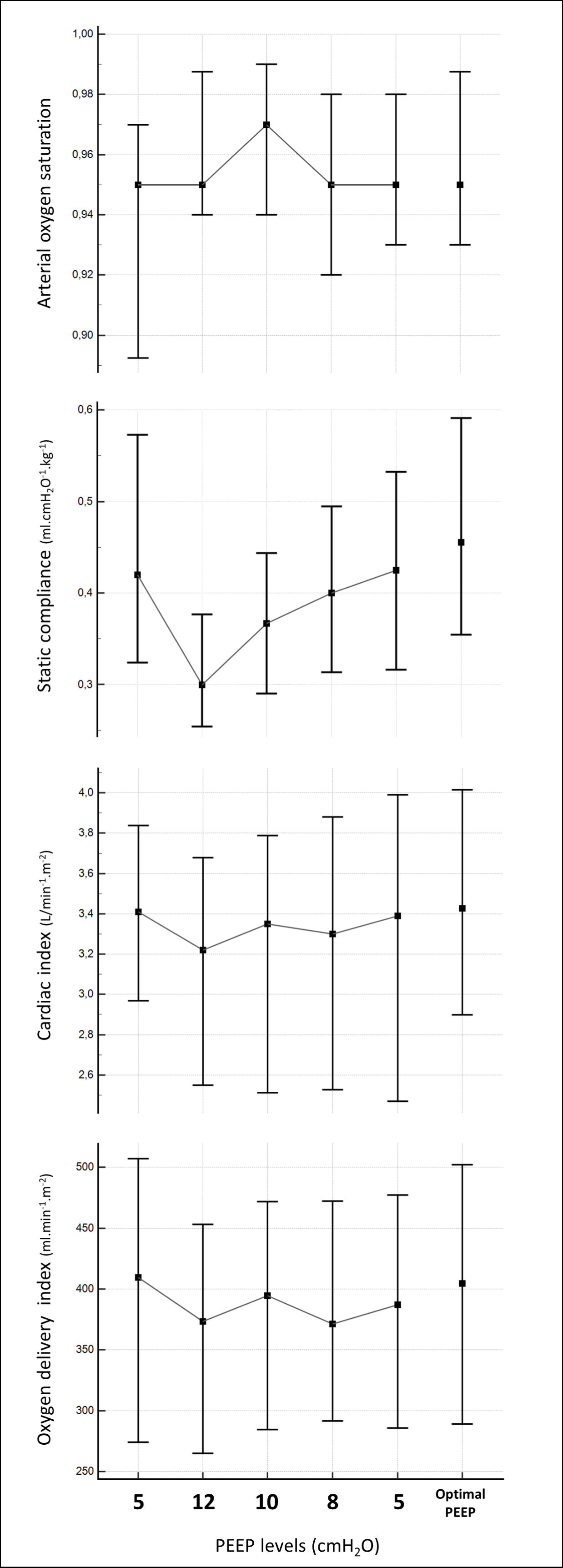
Arterial oxygen saturation, static compliance of the respiratory system, cardiac index, and estimated oxygen delivery index at different levels of positive end-expiratory pressure (PEEP) and optimal PEEP. The optimal PEEP was defined as the PEEP level where the best static compliance of the respiratory system was observed.

Additionally, we analyzed the Ci values obtained at the PEEP level where patients exhibited the highest static compliance, defined as the ’optimal PEEP’ (***Figure 1***). The optimal PEEP ranged from 5 to 10 cmH_2_O, with a median of 5 cmH_2_O (IQR 5-5). At the optimal PEEP, the median Ci was 3.43 L.min^-1^.m^-2^ (IQR 2.90 - 4.01), which was higher than the values observed at PEEP levels of 8, 10, and 12 cmH_2_O (*p*<0.05 for all), and similar to the values obtained at 5 cmH_2_O PEEP.

## DISCUSSION

Although it can improve oxygen saturation, inappropriately high PEEP levels have the potential to impact cardiac function and lung mechanics. In our study, we found significant reductions in stroke volume, cardiac output, and oxygen delivery when PEEP was increased to 12 cmH_2_O. Furthermore, the increase in PEEP was associated with a significant reduction in respiratory system compliance. However, it should be noted that the majority of participants in our study had characteristics that suggest a low potential for lung recruitability, such as mild or moderate PARDS with viral etiology. With this, our results reinforce the importance of closely monitoring hemodynamic variables and respiratory mechanics during PEEP titration in children with PARDS.

The PALICC-2 guidelines strongly recommend that PEEP levels should typically be maintained at or above the lower PEEP/higher FiO_2_ table from the ARDS Network protocol (14). This recommendation has the highest level of evidence present in the guidelines (moderate certainty of evidence), and was made based on studies that found increased mortality in patients with PARDS when PEEP levels below those recommended by the table were used (3, 4, 12). In addition, the authors recommend that practitioners perform a comprehensive assessment of hemodynamics, oxygenation/oxygen delivery, and static compliance of the respiratory system during PEEP titration (21). However, the evidence describing the adverse effects of PEEP in children is very scarce, especially involving PARDS patients managed with high PEEP levels.

Virk et al. measured the Ci of 15 children with mild to severe PARDS with a median PEEP of 8 cmH_2_O and subsequently with a PEEP of 4 cmH_2_O higher than baseline (22). No significant difference was observed before and after PEEP changes [4.4 L.min^-1^.m^-2^ (IQR 3.4 – 4.8) vs 4.3 L.min^-1^.m^-2^ (IQR 3.6 – 4.8), *p*=0.65]. The authors hypothesize that low lung compliance may prevent overdistension and thus attenuate the transmission of pressure from the airways to the pulmonary vascular bed. Some studies indeed suggest that the hemodynamic impact of PEEP is more pronounced when lung compliance is high (22–24). In pediatric swine models, the development of pulmonary overdistension by increasing tidal volume or PEEP was associated with a significant decrease in cardiac output (25). However, similar effects on cardiac function were seen in adults with acute respiratory distress syndrome (ARDS) and low lung compliance (9, 26, 27). So, the PEEP-induced cardiac output decrease may be more related to the occurrence of alveolar overdistention than to pulmonary compliance per se (6). In the present study, most of the patients included were admitted due to acute viral bronchiolitis and had mild or moderate PARDS. Therefore, it is very likely that the PEEP of 12 cmH_2_O was inappropriate, causing alveolar overdistension and thus reducing their cardiac output.

Few studies have evaluated the hemodynamic impact of PEEP in children. Ingaramo et al. performed hemodynamic evaluation in 50 children at PEEP levels of 0, 4, 8 and 12 cmH_2_O (23). Their results were similar to those herein reported. The median change in Ci observed was of 0.4 L.min^-1^.m^-2^ (< 10%) between the PEEP of 0 and 12 cmH_2_O. Although statistically significant, the authors considered this reduction to be minimal and, therefore, without clinical relevance. However, analysis of median values may not reveal significant changes that occurred in some individuals. In our study, while median values only reduced by about 5%, which could be considered clinically insignificant, 45% of participants experienced a greater than 10% reduction in SVi and Ci. Moreover, in line with our results, Ingaramo et al. also did not find significant changes in blood pressure when PEEP was increased. Thus, these findings emphasize that blood pressure should not be used as a surrogate for cardiac output during PEEP titration. It is noteworthy that this data was obtained from stable patients on relatively minimal ventilatory support and may not hold true for those with hemodynamic instability or PARDS.

Assessing the potential for lung recruitment is crucial to predict the benefits of PEEP and avoid its deleterious effects, including those on the cardiovascular system (28). Unfortunately, to discriminate consolidated and non-openable lung regions from those that are collapsed and recruitable for aeration may be a very difficult task. In adults with ARDS, recruitable regions may vary from 0% to 50% of the lung parenchyma (29). In our study, participants experienced reduced compliance of the respiratory system when increasing PEEP, which suggests low lung recruitability and alveolar overdistension. The aforementioned clinical characteristics of the included patients may partially explain this finding.

However, other pediatric studies evaluating children with PARDS also did not observe improvement in respiratory system compliance when increasing PEEP (22, 30). Virk et al. observed a statistically significant decrease in dynamic compliance from 0.38 ml.cmH_2_O^-1^.kg^-1^ (IQR 0.27 – 0.50) at a median PEEP of 8 cmH_2_O to 0.31 ml.cmH_2_O^-1^.kg^-1^ (IQR 0.22 – 0.42) when PEEP was increased by 4 cmH_2_O (22). In another study, respiratory system compliance of 15 children with PARDS remained unchanged between PEEP levels of 4 and 10 cmH_2_O (30). As in the present study, these authors evaluated respiratory mechanics after a few minutes of increasing PEEP, which may not have been enough time to significantly open the collapsed lung regions. Smallwood et al. conducted an evaluation on the time taken for a change in PEEP to exert its maximum impact on oxygenation and respiratory system compliance in children (31). They found that reaching 90% of the maximum observed change in dynamic compliance demanded 38 minutes, whereas optimal oxygenation required 71 minutes.

However, it is also possible that potential for alveolar recruitability is low when PARDS is caused by a primary lung disease, similar to what occurs in adults with ARDS (29). There seem to be substantial differences in respiratory mechanics between ARDS caused by pulmonary disease and ARDS originating from extrapulmonary diseases. Gattinoni et al observed that increasing PEEP led to opposite effects on respiratory mechanics in these two types of ARDS (29). Whereas patients with extrapulmonary ARDS experienced an increase in lung compliance, subjects with pulmonary ARDS showed a marked increase in lung elastance. Nevertheless, PEEP titration based only on respiratory mechanics may not be an appropriate method. Chiumello et al. found no association between lung mechanics-based variables and lung recruitability in adults with ARDS (32). It is important to emphasize that the results of these studies should not be extrapolated to PARDS, as the respiratory mechanics of children significantly differ from adults. Therefore, further studies evaluating lung recruitability in PARDS should take into account its severity, respiratory mechanics, hemodynamic status and etiology.

In the present study, participants experienced increased SatO_2_ at higher PEEP levels. PEEP titration based on oxygenation is probably the most popular method used at the bedside. Although several studies have shown that PEEP levels are not associated with survival in adults, a recent meta-analysis has shown that high PEEP was associated with reduced mortality in patients with ARDS who responded to increased PEEP by improved oxygenation (33). Interestingly, a prospective cohort study involving 352 children found that improvements in oxygenation (PaO_2_/FiO_2_ ratio) at 24 hours of PARDS onset, but not in respiratory mechanics, were associated with lower mortality (34). Thus, oxygenation must be strongly considered when setting up mechanical ventilation in children with PARDS.

Nevertheless, the effects of PEEP on oxygenation must be interpreted with caution, as alveolar recruitment is not the only mechanism responsible for the increase in arterial oxygen partial pressure (8). Due to the marked influence of cardiac function on the distribution of pulmonary blood flow, changes in cardiac output can substantially modify the degree of the intrapulmonary shunt. In the early study by Dantzker et al. involving 20 adults with ARDS, the application of 16 cmH_2_O PEEP led to a decrease in cardiac output from 9.0 to 4.5 L.min^-1^.m^-2^ (7). There was also a concomitant reduction in intrapulmonary shunt from 43.8% to 14.2%. Thus, the PEEP-induced decrease in cardiac output may play an important role in improving gas exchange. Several studies have observed a strong correlation between cardiac output and intrapulmonary shunt, such that reductions in cardiac output are usually associated with a reduction in the latter (7, 35, 36). However, despite the improvement in arterial oxygenation, the reduction in cardiac output can result in poor oxygen delivery, as observed in our study and previously by other authors (8, 9, 27). Therefore, it is critical that the interpretation of improvements in gas exchange during PEEP titration considers concomitant hemodynamic changes.

Some limitations of the present study need to be pointed out. First, in our study protocol, participants were assessed at specific PEEP values. Thus, it is not possible to draw conclusions about wider PEEP changes. Second, all participants were hemodynamically stable and most of them were admitted for acute viral bronchiolitis and had mild or moderate PARDS at enrollment. Although acute viral bronchiolitis is an important cause of PARDS, our results may not hold true for PARDS caused by other etiologies, for more severe cases, or for hemodynamically unstable patients (37). Third, Ci measurements were not performed using the accepted gold standard of thermodilution. However, Doppler cardiac output measurements have acceptable accuracy, precision, and repeatability in children (38). In addition, all echocardiographic measurements were performed by the same operator. Fourth, respiratory mechanics were not assessed using static measurements on volume-controlled ventilation mode and participants were not under neuromuscular blockade. Nevertheless, the analysis of respiratory system mechanics through the least squares fitting method has provided data that substantially agree with static methods and participants showed no signs of respiratory effort during the study protocol (39). Fifth, data were collected after a short period of 10 minutes at each PEEP level. Other results could have been found at the end of longer periods. Sixth, the operator was not blinded to the PEEP values during the echocardiographic examinations. Thus, operator bias was not prevented. Finally, the comparative analyzes do not take into account the precision of the devices used in this study.

## CONCLUSION

Although it may improve arterial oxygen saturation, increasing PEEP levels in PARDS patients with low potential for lung recruitability may reduce cardiac output, oxygen delivery, and respiratory system compliance. Therefore, due to the unpredictability of its adverse effects, PEEP titration should include close monitoring of cardiac output and respiratory system mechanics.

## Supporting information

Supplemental Digital Content

## Data Availability

All data produced in the present study are available upon reasonable request to the authors.

## Acknowledgments

Thank you to Carolina Grotta Ramos Telio for her review of the manuscript. We also thank the attending physicians, resident physicians, nursing staff, and legal guardians of the participants in this study.

