## Supplemental Digital Content for "The impact of PEEP on hemodynamics, respiratory mechanics, and oxygenation of children with PARDS"

**Funding Source:** No external funding for this manuscript.

**Financial Disclosure:** The authors have no financial relationships relevant to this study to disclose.

**Table of Contents**

[1.	Post-hoc analysis for pairwise comparison of data collected at different PEEP level according to Conover (1). Significance was defined as p < 0.05.	3](#_Toc143186881)

1. **Post-hoc analysis for pairwise comparison of data collected at different PEEP level according to Conover (1). Significance was defined as *p* < 0.05.**

### Variable: Heart rate

| Data collected at the PEEP level of: | Mean rank | Different (*p*<0.05) from variable number: |
| --- | --- | --- |
| (1) PEEP 5 cmH_2_O | 3.6452 | (4) (5) |
| (2) PEEP 12 cmH_2_O | 3.0968 | (5) |
| (3) PEEP 10 cmH_2_O | 3.2581 | (5) |
| (4) PEEP 8 cmH_2_O | 2.9032 | (1) (5) |
| (5) PEEP 5 cmH_2_O | 2.0968 | (1) (2) (3) (4) |

Minimum required difference of mean rank: 0.7225

### Variable: Cardiac index

| Data collected at the PEEP level of: | Mean rank | Different (*p*<0.05) from variable number: |
| --- | --- | --- |
| (1) PEEP 5 cmH_2_O | 3.6290 | (2) |
| (2) PEEP 12 cmH_2_O | 2.1129 | (1) (3) (4) (5) |
| (3) PEEP 10 cmH_2_O | 2.9194 | (2) |
| (4) PEEP 8 cmH_2_O | 2.9032 | (2) |
| (5) PEEP 5 cmH_2_O | 3.4355 | (2) |

Minimum required difference of mean rank: 0.7473

### Variable: Stroke volume index

| Data collected at the PEEP level of: | Mean rank | Different (*p*<0.05) from variable number: |
| --- | --- | --- |
| (1) PEEP 5 cmH_2_O | 3.6290 | (2) (3) |
| (2) PEEP 12 cmH_2_O | 1.9355 | (1) (4) (5) |
| (3) PEEP 10 cmH_2_O | 2.5484 | (1) (5) |
| (4) PEEP 8 cmH_2_O | 2.9516 | (2) (5) |
| (5) PEEP 5 cmH_2_O | 3.9355 | (2) (3) (4) |

Minimum required difference of mean rank: 0.6895

### Variable: Aortic velocity-time integral

| Data collected at the PEEP level of: | Mean rank | Different (*p*<0.05) from variable number: |
| --- | --- | --- |
| (1) PEEP 5 cmH_2_O | 3.6290 | (2) (3) |
| (2) PEEP 12 cmH_2_O | 1.9355 | (1) (4) (5) |
| (3) PEEP 10 cmH_2_O | 2.5484 | (1) (5) |
| (4) PEEP 8 cmH_2_O | 2.9516 | (2) (5) |
| (5) PEEP 5 cmH_2_O | 3.9355 | (2) (3) (4) |

Minimum required difference of mean rank: 0.6895

### Variable: Aortic blood flow peak velocity variation

| Data collected at the PEEP level of: | Mean rank | Different from: |
| --- | --- | --- |
| (1) PEEP 5 cmH_2_O | 3.2903 |  |
| (2) PEEP 12 cmH_2_O | 3.6290 | (3) (4) (5) |
| (3) PEEP 10 cmH_2_O | 2.7258 | (2) |
| (4) PEEP 8 cmH_2_O | 2.6452 | (2) |
| (5) PEEP 5 cmH_2_O | 2.7097 | (2) |

Minimum required difference of mean rank: 0.7596

### Variable: Pulmonary velocity-time integral

| Data collected at the PEEP level of: | Mean rank | Different from: |
| --- | --- | --- |
| (1) PEEP 5 cmH_2_O | 3.4839 | (2) (3) |
| (2) PEEP 12 cmH_2_O | 2.1290 | (1) (4) (5) |
| (3) PEEP 10 cmH_2_O | 2.5323 | (1) (5) |
| (4) PEEP 8 cmH_2_O | 3.1452 | (2) |
| (5) PEEP 5 cmH_2_O | 3.7097 | (2) (3) |

Minimum required difference of mean rank: 0.7261

### Variable: Peak inspiratory pressure

| Data collected at the PEEP level of: | Mean rank | Different from: nr |
| --- | --- | --- |
| (1) PEEP 5 cmH_2_O | 2.1452 | (2) (3) (4) (5) |
| (2) PEEP 12 cmH_2_O | 4.8387 | (1) (3) (4) (5) |
| (3) PEEP 10 cmH_2_O | 3.7097 | (1) (2) (4) (5) |
| (4) PEEP 8 cmH_2_O | 2.8226 | (1) (2) (3) (5) |
| (5) PEEP 5 cmH_2_O | 1.4839 | (1) (2) (3) (4) |

Minimum required difference of mean rank: 0.4073

### Variable: Mean airway pressure

| Data collected at the PEEP level of: | Mean rank | Different from: |
| --- | --- | --- |
| (1) PEEP 5 cmH_2_O | 1.9032 | (2) (3) (4) (5) |
| (2) PEEP 12 cmH_2_O | 4.8065 | (1) (3) (4) (5) |
| (3) PEEP 10 cmH_2_O | 3.9194 | (1) (2) (4) (5) |
| (4) PEEP 8 cmH_2_O | 2.9516 | (1) (2) (3) (5) |
| (5) PEEP 5 cmH_2_O | 1.4194 | (1) (2) (3) (4) |

Minimum required difference of mean rank: 0.3551

### Variable: Plateau pressure

| Data collected at the PEEP level of: | Mean rank | Different from: |
| --- | --- | --- |
| (1) PEEP 5 cmH_2_O | 1.7581 | (2) (3) (4) |
| (2) PEEP 12 cmH_2_O | 5.0000 | (1) (3) (4) (5) |
| (3) PEEP 10 cmH_2_O | 3.7742 | (1) (2) (4) (5) |
| (4) PEEP 8 cmH_2_O | 2.9194 | (1) (2) (3) (5) |
| (5) PEEP 5 cmH_2_O | 1.5484 | (2) (3) (4) |

Minimum required difference of mean rank: 0.3249

### Variable: Static compliance

| Data collected at the PEEP level of: | Mean rank | Different from: |
| --- | --- | --- |
| (1) PEEP 5 cmH_2_O | 3.5806 | (2) (3) |
| (2) PEEP 12 cmH_2_O | 1.4032 | (1) (3) (4) (5) |
| (3) PEEP 10 cmH_2_O | 2.5161 | (1) (2) (4) (5) |
| (4) PEEP 8 cmH_2_O | 3.6129 | (2) (3) |
| (5) PEEP 5 cmH_2_O | 3.8871 | (2) (3) |

Minimum required difference of mean rank: 0.5625

### Variable: Dynamic compliance

| Data collected at the PEEP level of: | Mean rank | Different from: |
| --- | --- | --- |
| (1) PEEP 5 cmH_2_O | 2.7581 | (2) (4) |
| (2) PEEP 12 cmH_2_O | 2.0000 | (1) (3) (4) (5) |
| (3) PEEP 10 cmH_2_O | 3.2097 | (2) |
| (4) PEEP 8 cmH_2_O | 3.7097 | (1) (2) |
| (5) PEEP 5 cmH_2_O | 3.3226 | (2) |

Minimum required difference of mean rank: 0.6863

### Variable: Inspiratory flow resistance

| Data collected at the PEEP level of: | Mean rank | Different from: |
| --- | --- | --- |
| (1) PEEP 5 cmH_2_O | 4.3226 | (2) (3) (4) |
| (2) PEEP 12 cmH_2_O | 1.6613 | (1) (4) (5) |
| (3) PEEP 10 cmH_2_O | 1.9677 | (1) (4) (5) |
| (4) PEEP 8 cmH_2_O | 2.9355 | (1) (2) (3) (5) |
| (5) PEEP 5 cmH_2_O | 4.1129 | (2) (3) (4) |

Minimum required difference of mean rank: 0.5095

### Variable: Inspired fraction of oxygen

| Data collected at the PEEP level of: | Mean rank | Different from: |
| --- | --- | --- |
| (1) PEEP 5 cmH_2_O | 3.3065 | (3) (4) (5) |
| (2) PEEP 12 cmH_2_O | 3.0806 |  |
| (3) PEEP 10 cmH_2_O | 2.8710 | (1) |
| (4) PEEP 8 cmH_2_O | 2.7903 | (1) |
| (5) PEEP 5 cmH_2_O | 2.9516 | (1) |

Minimum required difference of mean rank: 0.2973

### Variable: Peripheral oxygen saturation

| Data collected at the PEEP level of: | Mean rank | Different from: |
| --- | --- | --- |
| (1) PEEP 5 cmH_2_O | 2.3548 | (2) (3) |
| (2) PEEP 12 cmH_2_O | 3.1613 | (1) |
| (3) PEEP 10 cmH_2_O | 3.6290 | (1) (5) |
| (4) PEEP 8 cmH_2_O | 3.0323 |  |
| (5) PEEP 5 cmH_2_O | 2.8226 | (3) |

Minimum required difference of mean rank: 0.6833

### Variable: Oxygen saturation index

| Data collected at the PEEP level of: | Mean rank | Different from: |
| --- | --- | --- |
| (1) PEEP 5 cmH_2_O | 2.3710 | (2) (3) (5) |
| (2) PEEP 12 cmH_2_O | 4.7903 | (1) (3) (4) (5) |
| (3) PEEP 10 cmH_2_O | 3.7742 | (1) (2) (4) (5) |
| (4) PEEP 8 cmH_2_O | 2.7258 | (2) (3) (5) |
| (5) PEEP 5 cmH_2_O | 1.3387 | (1) (2) (3) (4) |

Minimum required difference of mean rank: 0.4377

### Variable: Estimated oxygen delivery index

| Data collected at the PEEP level of: | Mean rank | Different from: |
| --- | --- | --- |
| (1) PEEP 5 cmH_2_O | 3.7903 | (2) (3) (4) |
| (2) PEEP 12 cmH_2_O | 2.1290 | (1) (3) (5) |
| (3) PEEP 10 cmH_2_O | 3.0323 | (1) (2) |
| (4) PEEP 8 cmH_2_O | 2.8226 | (1) |
| (5) PEEP 5 cmH_2_O | 3.2258 | (2) |

Minimum required difference of mean rank: 0.7453

2. **Accuracy of devices used in the study**
2.1 **Mechanical ventilator - Hamilton-C1 (Hamilton Medical AG, Bonaduz, Switzerland)**

Accuracy of control settings and monitored parameters.

| **Control settings accuracy** | |
| --- | --- |
| PEEP (cmH2O) | ± 1cmH_2_O or ± 5%, whichever is greater |
| Oxygen (%) | ± (volume fraction of 2.5% + 2.5% gas level) |
| **Monitored parameters accuracy** | |
| Auto-PEEP (cmH2O) | ± 2cmH_2_O + 4% of actual reading |
| PEEP (cmH2O) | ± 2cmH_2_O + 4% of actual reading |
| Mean airway pressure (cmH2O) | ± 2cmH_2_O + 4% of actual reading |
| Peak inspiratory pressure (cmH2O) | ± 2cmH_2_O + 4% of actual reading |
| Plateau pressure (cmH2O) | ± 2cmH_2_O + 4% of actual reading |
| Expiratory tidal volume (ml) | ±10% or ±10 ml, whichever is greater |

The full description of accuracy is available *on the Hamilton Medical website: https://www.hamilton-medical.com/pt/Resource-center.html?ventilator=5db4d6d0-6fed-4b59-806e-edd19510d2cf&category=736b1b51-b3d2-4048-a64f-9bc268167da2&tab=d7dd4b8e-1047-4c29-a404-da2b41f2fa29&resource-lang=en&resource-detail-type=document&resource-detail-id=753ca119-aa50-4301-a2a9-45740b37cb1e*

*(see page 281-290)*

2.2 **Bedside monitor - Life Scope G5 monitors (CSM-1500; Nihon Kohden, Tokyo, Japan)**

Heart rate – precision of ± 2 bpm

Non-invasive arterial blood pressure – precision of ± 3 mmHg

Peripheral oxygen saturation – precision of ± 3%

The full description of precision is available at

<https://consultas.anvisa.gov.br/api/consulta/produtos/25351737421201411/anexo/T19765479/nomeArquivo/CSM%201900%201700%201500_4_Parte3.pdf?Authorization=Guest>

*(see chapter 17)*

2.3 **Ultrasound machine - GE Vivid q (GE Healthcare, Tirat Carmel, Israel)**

Spectral Doppler measurements – accuracy of 6%

The full description of precision is available at

[*https://customer-doc.cloud.gehealthcare.com/copyDoc/5407782-100/3*](https://customer-doc.cloud.gehealthcare.com/copyDoc/5407782-100/3)*).*

*(see page 63)*
